# Privacy-preserving large language model deployment for oncology registry abstraction: structure-aware evaluation in a real-world clinical setting

**DOI:** 10.64898/2026.05.18.26353541

**Authors:** Max Moldovan, Ruslan Enikeev, Megan Chu, Anisha Amalraj, Prajakta Prashant Koli, Shabbir Syed Abdul, Huren Sivaraj, Usman Iqbal, Chee Keong Toh

**Author notes:** Corresponding author: Dr. Max Moldovan, Registry of Senior Australians, South Australian Health and Medical Research Institute, Adelaide University, Adelaide, SA 5005, Australia.

## Abstract

**Objectives:** Structuring oncology clinical notes into registry-grade variables is essential for research and care but remains labour-intensive and error-prone. This study developed and evaluated a privacy-preserving large language model pipeline for oncology registry abstraction in a real-world clinical setting.

**Methods:** An open-source Meta Llama 3.3 70B-based pipeline, prompted without task-specific fine-tuning, was deployed to extract over 50 variables from 6,700 oncology notes at a cancer centre in Singapore. Data were de-identified locally using a Hide-In-Plain-Sight approach, ensuring no identifiable data left hospital infrastructure. Performance was assessed on 200 randomly sampled notes with adjudicated ground truth, using a structure-aware framework that classified outputs as correct, missing, spurious, or incorrect.

**Results:** F1 scores were high across variables, including diagnosis (97.2%), histology (95.8%), stage (92.6%), biomarkers (91.4%), and treatments (88.1%). Preliminary transferability testing on 50 external notes showed strong performance for core diagnostic variables, with reduced performance for list-valued variables.

**Discussion:** The structure-aware evaluation approach separated omissions from fabrications, providing more clinically actionable insight than aggregate metrics alone.

**Conclusion:** Privacy-preserving large language models achieved high extraction accuracy under a clinician-verified reference standard, supporting their feasibility for oncology registry abstraction in real-world clinical settings.

**What is already known on this topic:** Structuring oncology clinical notes into registry-grade variables is essential for cancer research and care but remains labour-intensive, costly, and inconsistent from one reviewer to the next when performed manually. Large language models offer potential to automate this task, but their use in real-world registries has been limited by concerns over patient data privacy, reliance on task-specific fine-tuning, and evaluation methods that obscure the difference between missing and fabricated information.

**What this study adds:** This study shows that an open-source, privacy-preserving large language model pipeline, deployed entirely within hospital infrastructure without task-specific fine-tuning, can extract over 50 registry variables from oncology notes with high accuracy (F1 scores 88–97% across key variables), validated against ground truth checked by two clinicians. Using a structure-aware evaluation framework, we demonstrate that errors can be meaningfully separated into missing, spurious, and incorrect categories, offering clearer insight into model behaviour than aggregate accuracy metrics alone, and show preliminary evidence of transferability to notes from an external setting.

**How this study might affect research, practice or policy:** These findings support the feasibility of deploying privacy-preserving, locally hosted large language models for oncology registry abstraction without requiring identifiable data to leave hospital systems or extensive model customisation. The structure-aware evaluation framework is also shared as a reusable template, enabling other institutions to validate similar AI extraction tools in a consistent, clinically meaningful way before wider implementation.

## Introduction

Oncology clinical notes are among the most challenging clinical documents to structure. Unlike radiology or pathology reports, which often follow semi-structured templates, oncology notes are narrative, longitudinal, and clinically dense, spanning diagnoses, treatment lines, biomarkers, staging, and clinical reasoning [1]. Abstracting these data into registry variables is traditionally slow, costly, and subject to inter-abstractor variability. Prior NLP studies have demonstrated feasibility but have generally focused on narrower tasks or specific endpoints [2]. As cancer centres increasingly depend on structured data for precision-oncology trials and real-world evidence, scalable abstraction is increasingly important.

Large language models (LLMs) offer a potential approach. Recent studies show that proprietary and open-source LLMs can extract structured information from clinical text with performance approaching dedicated NLP pipelines [3, 4]. Systematic reviews suggest that LLMs may offer advantages over traditional machine-learning classifiers for some healthcare text tasks [5], while domain-specific systems such as HARMON-E have achieved strong performance on oncology corpora [6]. Clinical deployment nevertheless raises privacy concerns, as patient notes contain protected health information subject to HIPAA, GDPR, and Singapore’s PDPA [7]. Federated learning is one approach [8-10], but requires multi-site infrastructure and coordination.

Data-localised deployment, where the LLM operates within hospital infrastructure and identifiable data remain within institutional boundaries, provides a simpler alternative. Although this approach has been explored [11], existing evaluations often use curated datasets, narrow document types, or metrics that obscure clinically important error modes, such as omissions versus hallucinations.

We report a real-world deployment of an open-source LLM pipeline for oncology registry abstraction at a private cancer centre in Singapore. The system generates the Well Characterized Repository (WCR), a continuously updated structured dataset containing over 50 oncology variables extracted from clinical notes and related documents using Meta Llama 3.3 70B without task-specific fine-tuning. We describe its data-localised, privacy-preserving architecture; introduce a structure-aware framework that evaluates binary, scalar, and list-valued fields using Correct, Missing, Spurious, and Incorrect categories [12]; and evaluate the framework on 200 independent notes with adjudicated ground truth, with bootstrap confidence intervals and exploratory transferability assessment on 50 notes from Taiwan.

## Materials and methods

### Ethics approval

This retrospective, non-interventional study evaluated a privacy-preserving large language model– based oncology registry abstraction system using de-identified clinical notes and related documents provided by Curie Oncology Pte Ltd, Singapore. The study team first accessed the de-identified data on 19 September 2025. No patients were recruited or contacted, no clinical intervention was performed, and clinical care was not altered. Curie Oncology, as the institutional data custodian, authorised access to and use of the de-identified data for development and refinement of the extraction pipeline, evaluation against clinician-reviewed reference data, and preparation of aggregated results for publication. Based on the nature and purpose of the activity, Curie Oncology determined that this retrospective, non-interventional evaluation did not constitute Human Biomedical Research within the scope of Section 3 of Singapore’s Human Biomedical Research Act 2015; accordingly, Institutional Review Board (IRB) review was not required. Data were processed under appropriate de-identification and access controls, and directly identifiable patient information was neither provided to the language model nor disclosed outside the authorised institutional environment. Patient-level clinical notes remain subject to institutional data-governance and confidentiality requirements and are not publicly available. Only aggregated findings, evaluation metrics, and non-identifiable results are reported.

### System architecture and privacy design

The pipeline operates within the hospital’s infrastructure. Clinical notes and laboratory reports are retrieved from the electronic medical record (EMR) system via secure application programming interface (API) connections. All preprocessing, including HTML flattening, field extraction, and de-identification, occurs on-premises. Before any data leaves the hospital network, personally identifiable information (PII) is masked using the Hide-In-Plain-Sight (HIPS) approach, which replaces sensitive entities (names, identifiers, dates of birth) with realistic but fictitious equivalents rather than generic placeholders. This preserves the linguistic structure of the note while removing re-identification risk. Only de-identified content is transmitted to a hospital cloud environment for LLM processing (Fig 1).

**Fig 1.**
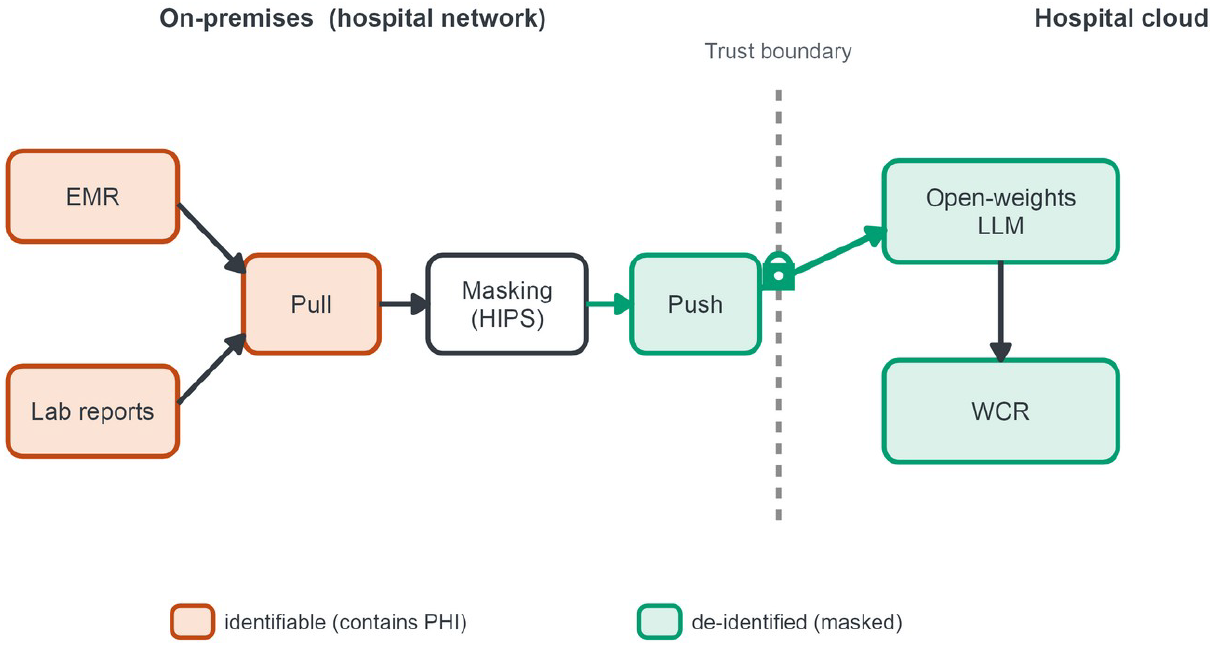
Privacy-preserving, data-localised deployment architecture for oncology registry abstraction.

The LLM is accessed via API within the hospital’s security boundary, and all identifiable data remain on-premises. The cloud infrastructure is defined as Infrastructure as Code (IaC), supporting deployment on AWS, Azure, GCP, or on-premises clusters. Access is restricted to secure API connections with no external internet exposure for database or processing components. The system operates within frameworks compliant with ISO 27001/17/18, SOC II, HIPAA, GDPR, and Singapore’s PDPA [16].

### Large language model configuration and prompting

The extraction engine is the Meta Llama 3.3 70B model, implemented as a multi-step, modular prompting pipeline. The model first collects domain-specific clinical evidence from the note across major oncology areas (diagnosis, staging, treatments, clinical status, and genomic findings). In subsequent steps, this evidence is classified and mapped into predefined registry fields using oncology-specific instructions and few-shot examples. The prompting is context-aware and designed to support temporal interpretation of longitudinal notes, including distinctions between historical and active disease, first versus second primary cancers, and sequencing of treatment lines. The pipeline also incorporates self-evaluation and confidence judgement to support downstream review workflows. No parameter fine-tuning was performed.

The LLM extracts clinically meaningful textual representations as expressed in the source note (e.g., “Triple Negative Breast Cancer”, “NSCLC”, “Carbo”) rather than standardised oncology codes. Downstream coding to ICD-10, ICD-O-3, or RxNorm is carried out separately, as a manual mapping step, and is not evaluated in this study. This design decouples extraction accuracy from terminology normalisation, allowing independent optimisation of each stage. No localised lookup tables, brand-name dictionaries, or site-specific gazetteers were used at any stage. Entity recognition and normalisation relied entirely on the native parametric knowledge of the Meta Llama 3.3 70B model.

### Dataset and sampling

From 6,700 clinical notes available at the hospital, we selected two non-overlapping subsets. While widely used benchmark datasets, such as MIMIC-III, have enabled reproducible research in critical care [13], they do not capture the complexity and longitudinal structure of oncology notes addressed in this study. A development set of 200 notes was purposively selected by the institution’s principal investigators to ensure broad coverage across major cancer types (breast, lung, colorectal, haematologic, and rare cancers), disease stages (early vs. advanced), and treatment modalities (surgery, chemotherapy, immunotherapy, targeted therapy). This set was used exclusively for iterative prompt development and output schema refinement. The prompt templates and output schema were then locked, and the test set was scored with that frozen version and never used to adjust prompts or schema.

Next, an independent test set of 200 clinical notes was randomly sampled from the remaining 6,500 notes with no overlap to the test set. All reported performance metrics are computed on this test set. While 200 notes represent approximately 3% of the available corpus, they yield reasonably tight bootstrap confidence intervals for the reported fields (Table 1), though the sample size still possesses a challenge for subgroup analyses by cancer type or disease stage. Expanding the test set is part of ongoing work.

**Table 1.** Extraction performance for 200 randomly sampled clinical notes.

| Field | Field-present | COR | INC | MIS | SPU | Precision | Recall | F1 | Specificity |
| --- | --- | --- | --- | --- | --- | --- | --- | --- | --- |
| First Primary Diagnosis | 158 | 154 | 3 | 1 | 2 | 96.9% (94.1–99.4) | 97.5% (94.8–99.4) | 97.2% (94.7–99.1) | 95.2% (87.8–100) |
| First Primary Histology | 130 | 124 | 5 | 1 | 0 | 96.1% (92.4–99.2) | 95.4% (91.4–98.5) | 95.8% (92.0–98.8) | 100% (100–100) |
| First Primary Overall Stage | 94 | 88 | 0 | 6 | 8 | 91.7% (85.7–96.8) | 93.6% (88.2–97.2) | 92.6% (88.4–96.2) | 92.5% (87.0–97.1) |
| Treatment Drugs | 103 | 352 | – | 40 | 55 | 86.5% (80.7–91.6) | 89.8% (83.2–95.4) | 88.1% (83.4–92.1) | – |
| Biomarker Test Results | 102 | 301 | – | 41 | 16 | 95.0% (90.3–98.4) | 88.0% (82.4–92.8) | 91.4% (87.3–94.7) | – |

### Ground truth and inter-annotator agreement

Ground-truth labels for the test set were established through dual independent clinical review followed by senior adjudication. Two clinical validators independently assessed LLM-extracted outputs against the source clinical notes for every case and field. Validators worked independently and without access to each other’s assessments. They were instructed to perform independent verification against the source note rather than accept model outputs at face value. This mirrors the intended real-world workflow, in which automated extraction is reviewed and verified by clinicians rather than annotated blind and de novo. The validators therefore assessed the model output against each source note rather than labelling independently of it, so the reference standard reflects clinician verification rather than blind labelling.

Inter-annotator agreement prior to adjudication was high. Using the same structure-aware evaluation framework as model assessment, agreement was computed bidirectionally (annotator A vs B and B vs A) and averaged. F1 ranged from 92.8% for list-valued variables (Treatment Drugs, Biomarker Test Results) to 96.8% for scalar diagnostic fields (Diagnosis, Histology), with disagreement primarily driven by missing and spurious extractions rather than incorrect values. In cases of disagreement, predominantly involving ambiguous staging or borderline drug classifications, a senior medical oncologist performed adjudication to establish the final reference standard. This inter-annotator agreement defines the clinician-verification reference standard for the task. Two trained reviewers, working independently, agree at an F1 of roughly 93 to 97%, which is the practically relevant comparator against which the model is assessed.

### Evaluation framework

For each field in each note, the LLM output was classified into one of four categories following MUC-style template-filling conventions developed in early information-extraction evaluations [12]:

- Correct (COR): output matches the ground truth.
- Missing (MIS): ground truth contains a value but the model output is empty or null.
- Spurious (SPU): the model outputs a value not present in the ground truth.
- Incorrect (INC): for scalar fields, the model outputs a value that differs from the ground truth.

For binary fields (e.g., presence or absence of metastasis), standard confusion-matrix elements (TP, TN, FP, FN) are computed, yielding accuracy, precision, recall, and specificity. For scalar fields (Diagnosis, Histology, Overall Stage) and list-valued fields (Treatment Drugs, Biomarker Test Results), metrics are computed from COR, INC, MIS, and SPU counts as follows (Fig 2).

**Fig 2.**
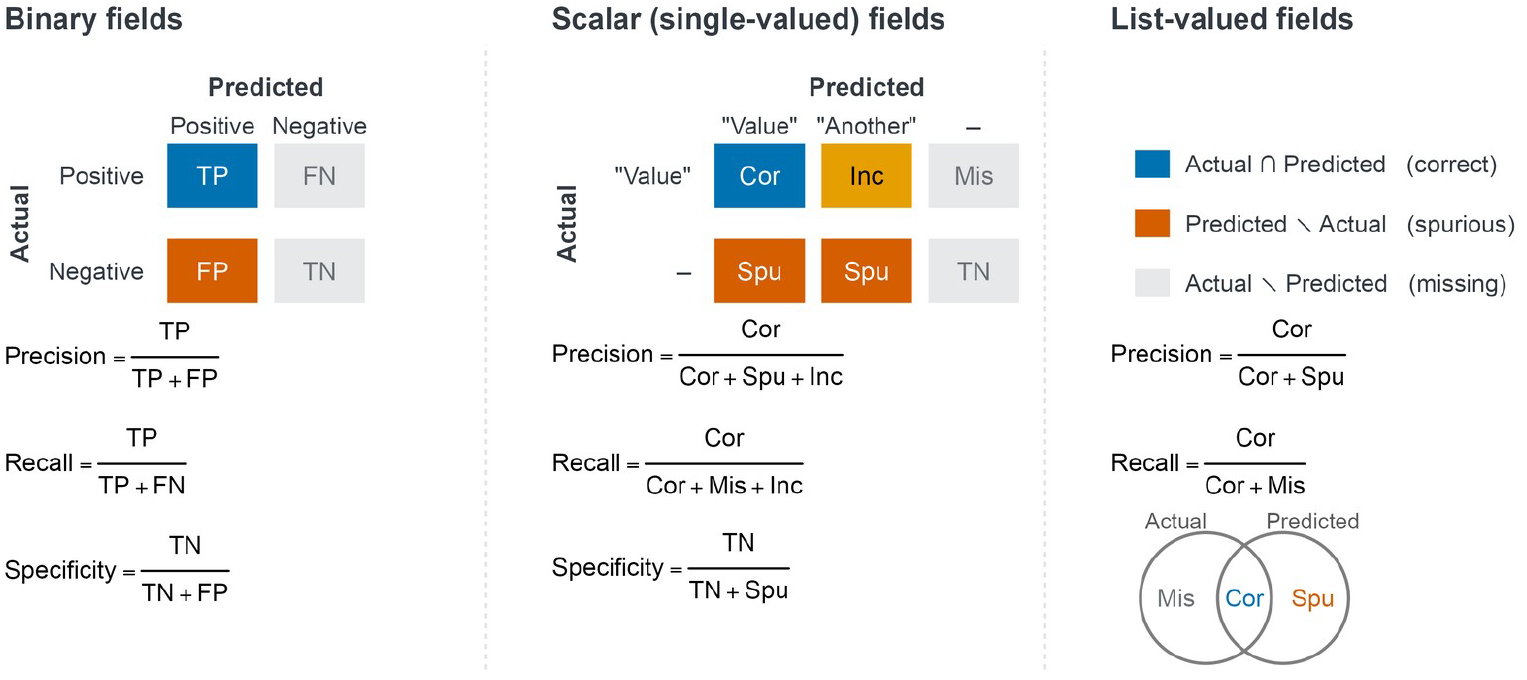
Structure-aware evaluation metrics for binary, scalar, and list-valued fields.

Scalar and binary fields are scored at the note level (a single value per note), whereas list-valued fields (Treatment Drugs, Biomarker Test Results) are scored at the item level, with each individual drug or biomarker result treated as a separate element. The COR, MIS, and SPU counts for list-valued fields consequently exceed the number of notes in which the field is present. This framework is structure-aware in two senses: it handles binary, scalar, and list-valued fields within a single evaluation run, and it separates omissions (MIS) from hallucinations (SPU) – a distinction that exact-match metrics collapse. Earlier evaluation approaches in clinical NLP tend to apply a single metric type uniformly [3, 12]. The mixed-type registry setting that oncology demands requires a more flexible scheme.

### Statistical methods

95% confidence intervals were computed using non-parametric bootstrap resampling at the note level, with B = 5,000 replicates [14]. For each replicate we resampled 200 notes with replacement, recomputed all field-level metrics, and reported the 2.5th and 97.5th percentiles as confidence bounds. Note-level resampling preserves the correlation structure among fields within a note.

## Results

Table 1 summarises extraction performance for five representative registry variables on the 200-note test set. These five are the core diagnostic and treatment variables that a cancer registry must capture (primary diagnosis, histology, overall stage, systemic treatments, and biomarker results), and they span both the scalar and list-valued field types that the framework scores. All performance estimates reflect comparison against adjudicated ground truth.

Core diagnostic variables showed excellent performance. First Primary Diagnosis achieved an F1 of 97.2% (95% CI 94.7 to 99.1), with errors almost entirely due to a small number of incorrect extractions (3 out of 158 field-present cases) rather than omissions or hallucinations. First Primary Histology was similarly strong (F1 95.8%, 92.0 to 98.8), with five incorrect values and perfect specificity, since the model never produced a histology output when none was warranted. Overall Stage had a lower but still high F1 of 92.6% (88.4 to 96.2), with errors split between missed stages (6 cases) and spurious outputs (8 cases), reflecting the inherent ambiguity of staging information scattered across longitudinal notes.

List-valued variables exhibited a different error profile. Treatment Drugs reached an F1 of 88.1% (83.4 to 92.1), with 40 missed items and 55 spurious extractions across 103 cases containing drug mentions. Biomarker Test Results achieved 91.4% (87.3 to 94.7), with better precision-recall balance (95.0% precision, 88.0% recall). When the model extracted a biomarker it was almost always correct, but it missed roughly one in eight results. Fig 3 shows per-field F1 with bootstrap confidence intervals against the inter-annotator agreement band, together with the decomposition of residual errors into missing and spurious modes.

**Fig 3.**
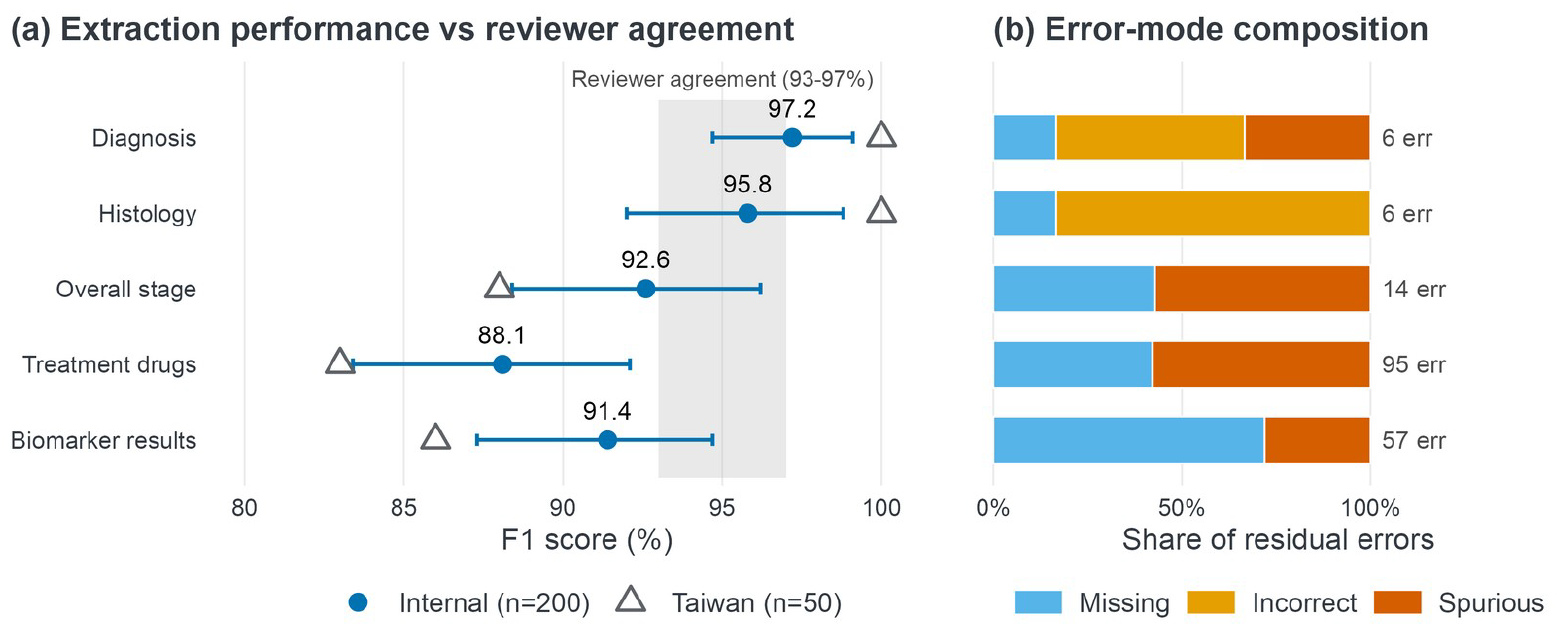
Per-field extraction performance with 95% confidence intervals and error-mode decomposition.

### Exploratory transferability assessment

To assess transferability, we applied the same pipeline, without modification and using the same prompt template, to 50 oncology notes from a large private hospital in Taiwan. F1 scores remained high for core diagnostic fields (Diagnosis 100%, Histology 100%), but were lower for complex variables: Overall Stage 88%, Treatment Drugs 83%, and Biomarker Test Results 86%. The drop was most pronounced for drug names, likely reflecting differences in prescribing conventions and brand-name usage between Singapore and Taiwan. These results suggest that the framework transfers across institutions, but that list-valued variables may benefit from site-specific prompt adaptation. As this external set comprises only 50 notes, the figures are point estimates reported without confidence intervals and should be read as an exploratory transferability signal rather than a precise generalisability estimate. Robust cross-site validation would require substantially larger external cohorts spanning multiple institutions and documentation styles, with per-field confidence intervals, which we regard as a priority for future work.

## Discussion

The results show that a general-purpose, open-source LLM can support registry-grade oncology data abstraction in a production clinical environment. F1 scores above 88% across all evaluated variables, and above 92% for core diagnostic fields, place this system within the performance range reported by recent dedicated extractors [4, 6]. Model performance approached the agreement level of two clinicians verifying extractions on the same workflow. As two independent human reviewers agreed at an F1 of 93 to 97% on this verification task, the model performs at the level of clinician verification rather than reaching a blind-abstraction ceiling. The operative comparator for a deployment study of this kind is, therefore, the manual abstraction workflow that the pipeline augments, not an alternative model architecture. The model meets that comparator for core fields while remaining consistent with systematic-review evidence that LLMs match or exceed traditional classifiers on clinical extraction tasks [5].

We deliberately prompted a general-purpose open-source model rather than fine-tuning a smaller task-specific one. Domain-specific and fine-tuned smaller models can match or exceed large general-purpose LLMs on well-defined, static extraction tasks, and where a fixed set of variables must be extracted at the highest possible accuracy, that approach is reasonable. However, for a production registry it is normally not the case. The set of required variables is not fixed, with new research questions and reporting requirements adding fields over time. A prompted model accommodates them by extending instructions, whereas a fine-tuned extractor would require fresh labelled data and re-training for every schema change. For sustained real-world deployment, where the schema evolves and labelling capacity is the binding constraint, such flexibility, rather than peak accuracy on a static variable set, is the operational requirement. For the same reason, a same-data comparison against a separately trained classifier, though a natural benchmarking exercise, would answer a different question from the one our deployment study asks. We, therefore, note it as future work, rather than a gap, in the present report.

Two aspects of these results deserve closer attention. First, the error-mode decomposition revealed that under-extraction (MIS), not hallucination (SPU), is the dominant failure mode. For Treatment Drugs, 42% of errors were missed items versus 58% spurious. For Biomarker Test Results the ratio reversed, with 72% missed and 28% spurious. This asymmetry has practical implications. In a human-in-the-loop workflow, a system that primarily under-extracts (MIS) would require reviewers to check for completeness, while one that over-extracts (SPU) demands scrutiny for false positives. Reporting these modes separately, rather than folding them into a single metrics, gives deployment teams the information needed to design appropriate review protocols.

Second, the transferability results from Taiwan are encouraging but not without limitations. Perfect F1 on Diagnosis and Histology suggests that core cancer-type recognition generalises well across sites. The performance drops for drugs and biomarkers, however, point to the expected challenge of institutional variation in clinical terminology, a well-documented issue in clinical NLP [1], which the pipeline did not mitigate with any localised dictionary. We regard site-specific prompt adaptation for list-valued variables as a practical next step rather than a fundamental limitation.

Liu et al. [15] documented the growing role of NLP for populating clinical registries. The evaluation framework presented here addresses a key methodological gap in this setting. By supporting binary, scalar, and list-valued fields within a unified scoring procedure, and by explicitly distinguishing omissions from hallucinations, it provides more actionable performance profiles than commonly used field-level exact-match or document-level correctness metrics, which collapse clinically important error modes. Unlike generic text-generation metrics or span-based evaluation approaches, the framework operates directly on structured outputs and decomposes errors into clinically interpretable categories. This is particularly important in oncology registry abstraction, where heterogeneous variable types must be evaluated within a single pipeline. The open-source release of the scoring code is intended to facilitate adoption and enable consistent comparison across sites.

Several limitations should be noted. First, the test set of 200 notes, while adequate for detecting large performance differences, limits statistical power for subgroup analyses by cancer type or disease stage. The 6,700-note corpus would support a larger evaluation, and we are expanding the test set in ongoing work. Second, all primary-site notes originated from a single English-speaking cancer centre in Singapore. Generalisability to multilingual environments, public hospitals with different documentation practices, or resource-limited settings remains unverified. The Taiwan pilot is a partial step, but 50 notes are insufficient for firm conclusions. Third, we evaluated text-level extraction accuracy, not downstream coding accuracy. Mapping extracted values to ICD-10 or RxNorm codes, a separate manual step, introduces additional error that we did not measure. Fourth, while the HIPS masking approach preserves linguistic structure, it could introduce artefacts if entity boundaries are misidentified, a risk we did not formally quantify. Finally, the reference standard was established by clinician verification rather than blind annotation. Both validators reviewed the model output against each source note rather than labelling independently of it, so the ground truth, and the inter-annotator agreement derived from it, reflect a verification workflow rather than blind de novo labelling. This matches how the system is used in practice, where clinicians review automated extractions, but it can make the performance estimates optimistic relative to fully blinded annotation, and the results should be read in that light.

## Conclusion

We presented a privacy-preserving LLM pipeline for oncology registry abstraction, deployed in a real clinical setting and evaluated with structure-aware metrics on adjudicated ground truth. The results confirm that prompted open-source models can achieve registry-grade extraction quality without fine-tuning, and that error-mode decomposition provides actionable insight beyond aggregate accuracy scores. The open-source evaluation framework supports transparent comparison across institutions. Future work will expand the evaluation to multi-site deployments, larger and more diverse test sets, and formal comparison of abstraction costs against manual workflows.

## Data Availability

Patient-level clinical notes cannot be shared due to privacy and institutional restrictions

https://github.com/Oncoshot/llm-validation-framework

## Code availability

The evaluation and validation framework used in this study is available as open-source software at https://github.com/Oncoshot/llm-validation-framework. Patient-level clinical notes cannot be shared due to privacy and institutional restrictions. The released code enables replication of the evaluation methodology on local datasets. Infrastructure deployment specifications are described in a companion technical report [16].

## Acknowledgments

The authors thank the clinical validators who established the ground-truth reference standard, and Curie Oncology Pte Ltd for institutional data-governance support of this evaluation.

